# From Symptom Networks to Conversation Networks: A Cross-Sectional Study Mapping the Topology of Suicide-Related Clinical Dialogue

**DOI:** 10.64898/2026.09.27.26364136

**Authors:** Oliver Grimm, Christopher Landau, Sofia Arampatzi, Anmol Goel, Eugen Owtscharow, Hiba Arnaout, Iryna Gurevych, Andreas Reif

## Abstract

Suicide risk assessment in clinical conversations relies on the sequential structure of therapist-patient dialogue, yet formal models of how suicide-related content emerges, transitions, and reciprocates across conversational turns are lacking. We applied network analysis to C-SSRS-coded utterances from psychotherapy sessions to characterize the topology and dynamics of suicide-related clinical communication. Transcripts of 110 psychotherapy sessions from the SPEAK-SAFE study were classified for suicidality relevance using the Qwen3-32B large language model with an adapted Columbia-Suicide Severity Rating Scale (C-SSRS) schema. From the suicidality-relevant utterances, we constructed a weighted directed category transition network. Centrality, reciprocity, clustering, assortativity, and path length metrics were computed. Independent human expert ratings for a session-level subsample of 30 sessions from this cohort showed substantial agreement with the LLM classifications (Spearman ρ = 0.885, ICC[2,1] = 0.708). The network was dominated by therapist inquiry. Reciprocity was high, and closeness centrality decreased monotonically with C-SSRS severity. Active suicidal ideation functioned as a bridge node from which escalation pathways to method, plan, and attempt categories originated. The network exhibited a hub-and-spoke topology with small-world characteristics. Suicide-related clinical dialogue is structured as a therapist-centered hub-and-spoke network with high reciprocity and a consistent severity-peripherality gradient. The extension of network theory from intra-individual symptom structures to inter-individual communication dynamics represents a methodological innovation demonstrated here as a proof of concept, with potential applications to clinical training, automated decision support, and multimodal risk assessment. clinical interviews clinical interviews

**Author Summary:** clinical interviews clinical interviewsWhen clinicians talk with patients about suicide risk, the conversation itself follows a pattern: the therapist asks, the patient answers, and the topic shifts from one aspect of risk to another. We wanted to know whether this back-and-forth has a describable structure, in the same way that networks of interacting symptoms have been used to describe mental disorders. Using a large language model, we automatically labeled which parts of 110 real psychotherapy conversations touched on suicidal thoughts, plans, or behaviors, and then built a network showing how the conversation moved between these topics. We found that the therapist’s questions sat at the center of this network, that the conversation frequently looped back to earlier topics, and that mentions of active suicidal thoughts acted as a gateway to discussion of more severe risk, such as specific plans or past attempts. This proof-of-concept approach offers a new way to visualize and quantify how suicide risk assessments unfold in real clinical dialogue, with potential future applications in clinician training and automated support tools.

## Introduction

Suicide remains one of the leading causes of death among individuals with psychiatric disorders, accounting for approximately 700,000 deaths annually worldwide [1]. In clinical practice, the assessment and management of suicide risk is among the most demanding tasks for mental health professionals, and one for which many healthcare professionals report feeling inadequately prepared. A recent qualitative study found that 85% of community mental health clinicians spontaneously reported anxiety or heightened emotional distress when discussing their suicide prevention practices [2]. Clinicians consistently described low self-efficacy in suicide prevention skills and difficulty tolerating the uncertainty of risk assessment. This anxiety has measurable consequences: clinician confidence in suicide risk decision-making is negatively associated with psychological stress, physiological stress, and emotional exhaustion [3].

(e.g. how do topics transition) s?HThe therapeutic conversation is the usual vehicle for suicide risk assessment, yet its sequential structure remains understudied. A systematic review of 37 studies characterized the therapeutic alliance with suicidal clients as “working on the edge” and found that flexibly interweaving risk assessment into the natural flow of conversation strengthens the therapeutic relationship [4]. This finding raises an empirical question with clinical relevance: what is the natural structure of suicide-related clinical dialogue, and how can formal modeling of that structure generate clinical insights?

TOver the past decade, the network theory of mental disorders [5,6] has fundamentally reframed how psychopathology is conceptualized. Rather than treating mental disorders as the expression of latent disease entities, the network approach posits that disorders are their symptom networks: patterns of mutually reinforcing causal interactions between symptoms. This framework has been empirically operationalized through cross-sectional network models in which nodes represent symptoms and edges represent partial correlations, with centrality metrics identifying the most structurally influential symptoms as intervention targets [7]. In the specific domain of suicidality, symptom-level network analyses have identified difficulty identifying feelings and personal distress as central bridge nodes connecting emotional dysfunction to suicidal ideation in adolescents with major depressive disorder [8].

A critical theoretical gap separates intra-individual symptom networks—where nodes represent symptoms and edges represent cross-sectional partial correlations—from the inter-individual communication networks through which suicidality is assessed. In such a communication network, nodes represent clinically meaningful utterance categories (here, C-SSRS codes assigned during the dialogue), and directed edges represent temporally ordered transitions between categories as the conversation unfolds. The clinical interview is a dyadic interaction in which the therapist actively probes, the patient selectively discloses, and the sequential dynamics of the exchange shape what is revealed and what remains hidden [4]. Recent work by de Felice et al. [9] applied Markov transition networks to model moment-to-moment shifts between abstract and emotional language states within psychotherapy sessions, demonstrating the feasibility of network analysis for therapeutic dialogue—but their node definitions remained at the level of broad linguistic modes rather than clinically grounded suicide risk categories. Existing network-based approaches to clinical communication have either modeled who speaks to whom in multidisciplinary team meetings without modeling utterance content [10] or used static semantic similarity networks to predict disclosure of self-harm ideation without characterizing the temporal transition dynamics themselves [11]. Thus, no prior study has applied the full suite of directed graph metrics to the sequential category structure of suicide risk assessment dialogue.

The present study applies network analysis to the sequential category structure of 110 face-to-face clinical interviews from the SPEAK-SAFE study [12]. We propose an extension of Borsboom’s network-theoretic framework from intra-individual symptom structures to inter-individual communication dynamics: C-SSRS categories serve as nodes, and their temporally adjacent transitions within clinical dialogue serve as directed edges. A transition is defined as a consecutive pair of suicidality-relevant utterances (c_p, c_q) belonging to distinct C-SSRS categories. For example, a therapist inquiry about suicidal ideation immediately followed by a patient disclosing active ideation constitutes a THERAPIST_INQUIRY → ACTIVE_IDEATION transition. Self-transitions (e.g., a sustained sequence of therapist probes) are excluded from the graph. They carry no information about category switching behavior and would otherwise dominate the network. Transitions are aggregated across all 110 sessions: each observed transition increments an edge weight w_{p,q}, producing a weighted directed graph in which higher edge weights identify clinically common conversational pathways already visible in the natural flow of the intake interview. The therapist was modeled as a distinct node (THERAPIST_INQUIRY) rather than being merged with patient categories, because therapist utterances are probing interventions rather than symptom disclosures; retaining the therapist as a separate node preserves the directional structure of clinical elicitation, which is essential for testing hypotheses about how therapist-initiated transitions differ from patient-initiated ones. The methodological workflow is illustrated in Fig 1.

**Fig 1.**
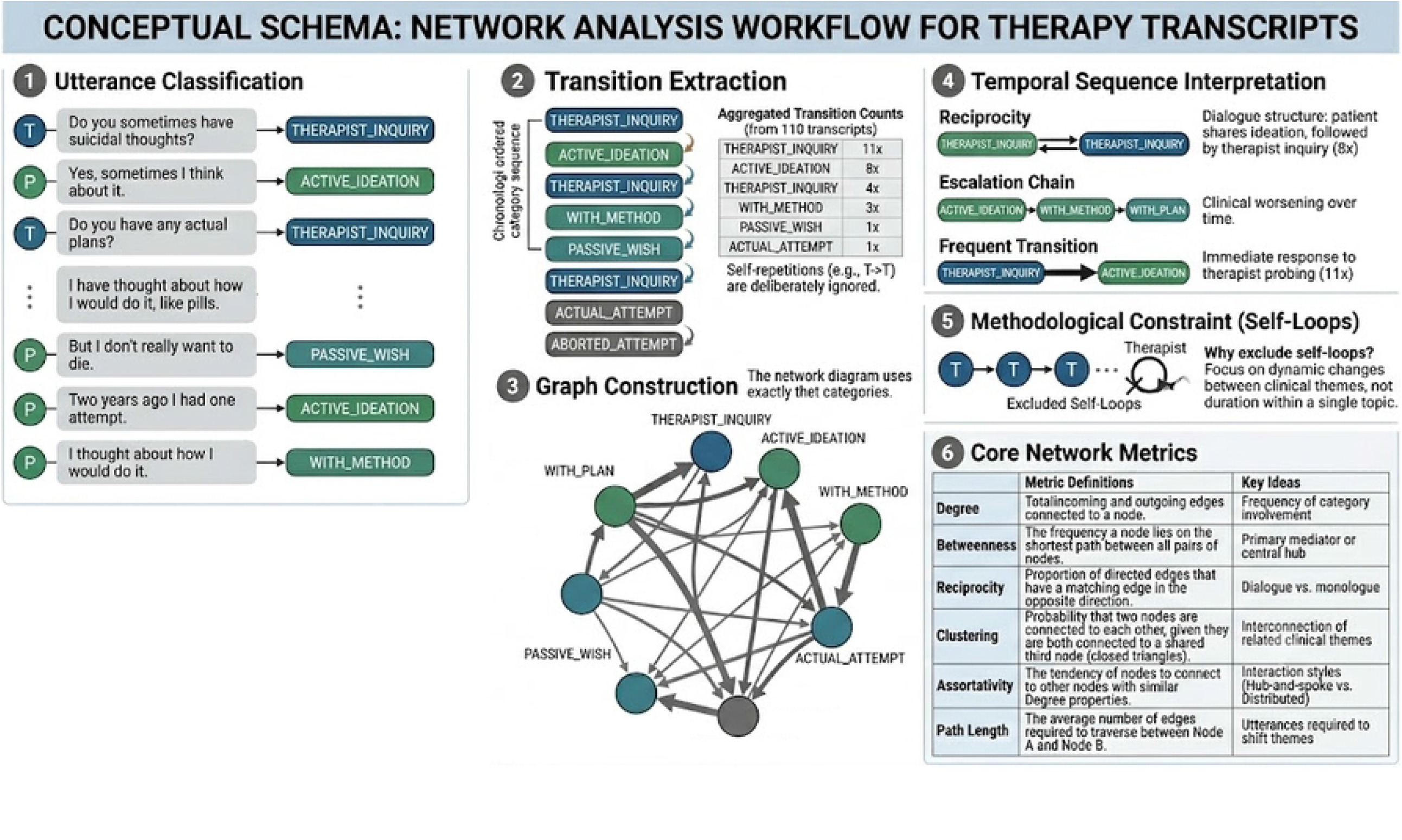
Conceptual Schema for the Category Transition Network Analysis of Suicidality-Related Clinical Dialogue. The figure details the six-step method for converting raw therapeutic dialogue into structured, quantifiable network data. Step 1 (Utterance Classification): Raw sequential dialogue from therapists (T) and patients (P) is automatically categorized into C-SSRS labels using Qwen3-32B. Step 2 (Transition Extraction): Chronologically adjacent utterances are parsed to identify categorical shifts; self-repeating categories are excluded to isolate dynamic theme changes. Transition frequencies are aggregated across all 110 transcripts (96 total transitions, 32 directed category pairs, 8 distinct categories). Step 3 (Graph Construction): A directed weighted graph G = (V, E, w) is built; edge thickness is proportional to transition frequency (range: 1–11), and arrow direction indicates temporal order. Step 4 (Temporal Sequence Interpretation): Specific structures are analyzed for clinical meaning, including reciprocity (R = 0.81, dialogical structure), escalation chains (e.g., ACTIVE_IDEATION → WITH_METHOD → WITH_PLAN), and dominant conversational paths (e.g., THERAPIST_INQUIRY → ACTIVE_IDEATION, w = 11). Step 5 (Methodological Constraint): Self-loops (**c_p** = **c_q**) are excluded to prioritize dynamic category shifts and prevent duration-based noise. Step 6 (Core Network Metrics): Six topological metrics are computed on the weighted directed graph; temporal sequence is implicitly encoded in all measures because they are derived from the directed edges representing aggregated temporally ordered transitions across all 110 sessions.

We tested four hypotheses. Hypothesis 1 (Hub-and-Spoke Topology) comprised three sub-predictions: (H1a) the therapist node would dominate all centrality metrics (in-degree, out-degree, betweenness, closeness), consistent with a therapist-centered hub; (H1b) the network would exhibit high dialogical reciprocity (R > 0.70), reflecting the turn-taking structure of clinical elicitation; and (H1c) closeness centrality would decrease monotonically with C-SSRS severity (severity-peripherality gradient), with more severe categories occupying more peripheral topological positions. Hypothesis 2 (Bridge Function of Active Ideation) comprised two sub-predictions: (H2a) ACTIVE_IDEATION would show the highest HITS authority score among patient categories, reflecting its role as the most frequent destination of therapist probes; and (H2b) ACTIVE_IDEATION would serve as a bridge node from which escalation pathways to higher-severity categories (WITH_METHOD, WITH_PLAN, WITH_INTENT, ACTUAL_ATTEMPT) originate. Hypothesis 3 (Severity-Avoidance Gradient) predicted that therapist-to-patient transition frequencies would be negatively correlated with the C-SSRS severity of the target patient category, such that therapists would transition less frequently to more severe content. Hypothesis 4 (Reciprocity–Exploration Association) predicted that sessions with higher dialogical reciprocity would explore a greater breadth of distinct C-SSRS categories, reflecting a more thorough clinical assessment. Given the anticipated sparsity of suicide-related content in a general psychiatric intake sample, we frame this investigation as a proof-of-concept demonstration of a novel methodological framework.

## Methods

### Participants and Data Source

The analysis draws on 110 clinical intake interviews (t1) from the SPEAK-SAFE study, a multicentric longitudinal investigation of adults undergoing psychiatric-psychotherapeutic treatment at the University Hospital Frankfurt and the Privatklinik Dr. Amelung, Germany [12]. The study was approved by the institutional ethics committee and registered at the German Clinical Trials Register (DRKS00027878). Participants were aged 18 to 65 years with ICD-10 diagnoses including schizophrenia spectrum disorders (F20.x, F23, F25) and affective disorders (F30–F33). All participants provided written informed consent. Interviews were conducted in German by trained clinicians between November 1, 2023, and [DATE: confirm data collection end date or ‘ongoing’], audio-recorded using Tascam DR-40X recorders with two condenser microphones (48 kHz, 24-bit WAV), and transcribed using WhisperX with subsequent manual pseudonymization. Transcripts were diarized into speaker-annotated utterances following the WebVTT standard, yielding a corpus of 55,796 utterance-level segments. Mean interview duration was 42.8 minutes (SD = 14.0, median = 46.9, range: 2.3–60.0). Interviews were conducted by a team of trained clinicians across both study sites (University Hospital Frankfurt and Privatklinik Dr. Amelung).

### Per-Utterance Suicidality Classification

Following the workflow illustrated in Fig 1 (Step 1), each utterance was independently classified for suicidality relevance using the Qwen3-32B large language model [13] with temperature T = 0 to ensure deterministic output. The model was deployed locally via LM Studio on an two NVIDIA RTX A6000 GPUs (2*48 GB VRAM), ensuring that no patient data left the institutional network. This privacy-preserving local deployment strategy was part of a detailed data security plan and workflow (see [12]). Classification was performed using an adapted Columbia-Suicide Severity Rating Scale (C-SSRS; [14]) schema comprising 11 patient-facing categories (PASSIVE_WISH, ACTIVE_IDEATION, WITH_METHOD, WITH_INTENT, WITH_PLAN, ACTUAL_ATTEMPT, INTERRUPTED_ATTEMPT, ABORTED_ATTEMPT, PREP_BEHAVIOR, non-suicidal self-injury (NSSI), THIRD_PERSON_MENTION) plus one therapist-specific category (THERAPIST_INQUIRY). Each utterance was presented to the model together with the preceding K = 8 utterances as a sliding context window, enabling disambiguation of third-person references and negations. The model returned, for each utterance, a binary flag indicating relevance for suicidality and a set of applicable C-SSRS categories. As depicted in Step 1 of Fig 1, the output is a chronologically ordered sequence of categorized utterances that serves as the input to the transition extraction step.

As independent evidence for the reliability of Qwen3-32B in this clinical population and language, a validation study compared session-level LLM ratings (C-SSRS ideation level and overall severity, generated by the same model deployed with the same prompting framework) against independent human expert ratings for a subsample of n = 30 t1 transcripts from this cohort. Agreement was substantial (overall severity: Spearman ρ = 0.885, ICC[2,1] = 0.708, quadratic-weighted κ = 0.70; 90.0% of ratings agreed within one severity level), and no case rated as suicidal by the human expert was rated as non-suicidal by the LLM. The LLM’s error pattern was a tendency toward slight overestimation (bias = +0.57). Independent rater validation was conducted at the whole-session level, for practical reasons, as utterance-level validation with >50,000 utterances was not possible for human raters; see Limitations.

### Transition Extraction

As shown in Fig 1, Step 2, the chronological sequence of categorized utterances is parsed to identify immediate categorical shifts between temporally adjacent utterances. For each session s, the set of differential transitions is defined as T(s) = {(c_p, c_q) : an utterance with category c_p immediately precedes an utterance with category c_q, and c_p ≠ c_q}. Self-transitions, in which the same category appears in consecutive utterances, are excluded because they carry no information about category switching behavior. Transition frequencies were aggregated across all 110 transcripts, yielding a global multiset of 96 transitions distributed across 32 directed category pairs involving 8 distinct C-SSRS categories.

### Graph Construction

Following the schema of Fig 1, Step 3, the aggregated transition frequencies are used to construct a directed, weighted graph G = (V, E, w), where V represents the set of 8 observed C-SSRS categories, each edge (u, v) ∈ E represents the existence of at least one observed transition from category u to category v, and the weight w(u, v) equals the absolute frequency of that transition across all sessions. Edge thickness in the visualization (Fig 2) is proportional to weight (range: 1–11). Arrow direction indicates temporal order. Node size is proportional to weighted total degree centrality. The resulting graph contained 8 vertices and 32 directed edges.

**Fig 2.**
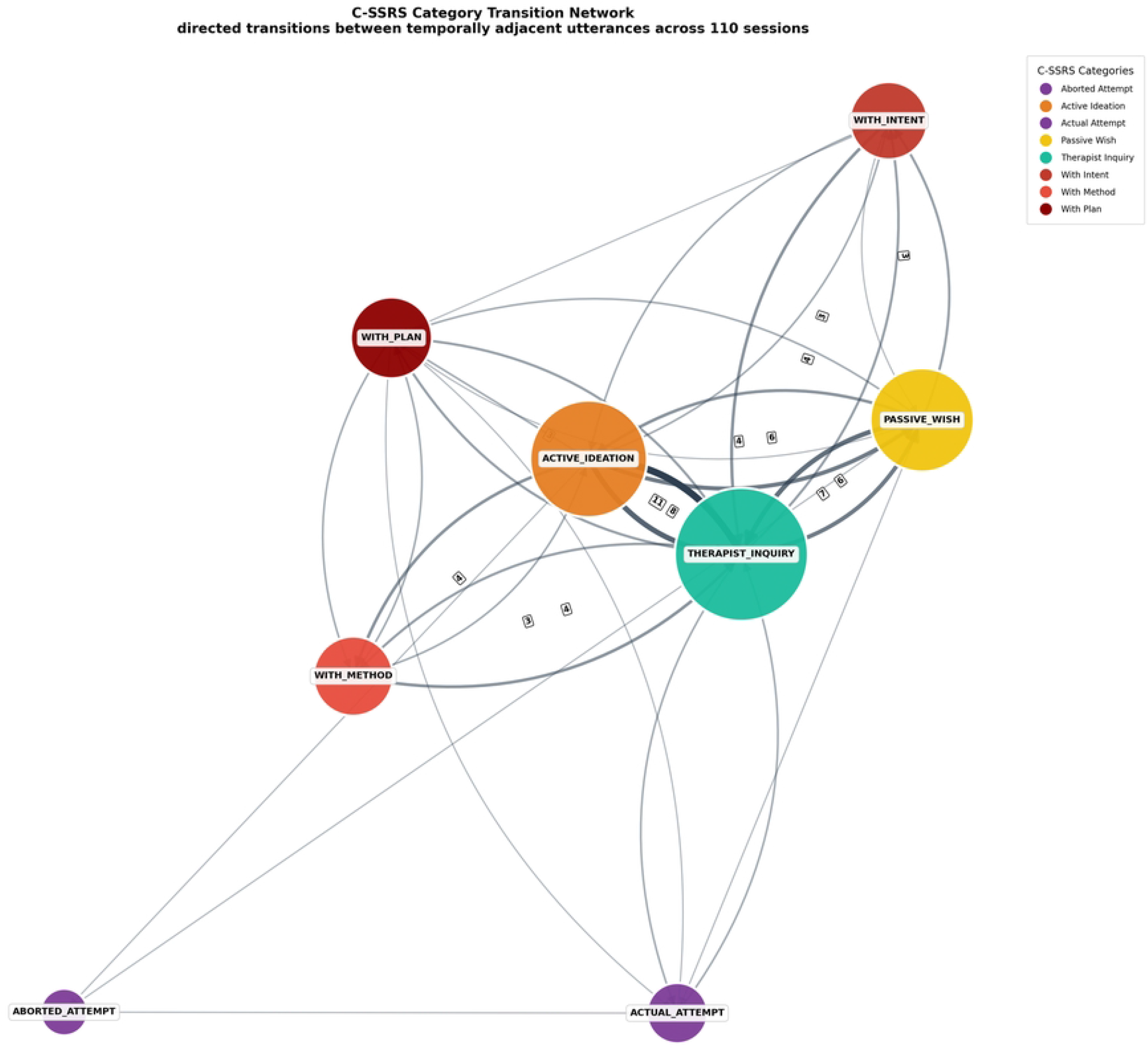
Category Transition Network of Suicide-Related Clinical Dialogue. Nodes represent C-SSRS categories assigned to the 506 suicidality-relevant utterances across 110 clinical intake interviews. Directed edges represent temporally adjacent transitions between categories, aggregated across all sessions; edge thickness and opacity are proportional to transition frequency (w; range: 1–11). Self-transitions are excluded. Node size is proportional to weighted total degree centrality; node color follows a clinical severity gradient from low (yellow: PASSIVE_WISH) through intermediate (orange: ACTIVE_IDEATION) to high (dark red: WITH_PLAN, ACTUAL_ATTEMPT), with therapist inquiries rendered in teal. Edge labels display transition frequency for edges with w ≥ 3. The layout was computed using the Fruchterman-Reingold force-directed algorithm [24]. An interactive Cytoscape.js version [25,26] is available in the online supplement.

### Network Metrics and Temporal Interpretation

Consistent with the analytic framework depicted in Fig 1, Step 6, the following network metrics were computed. Weighted in-degree, out-degree, and total degree quantified the flow of transitions through each category [15]. Betweenness centrality C_B identified categories serving as bridging points on shortest paths between other categories [16].

Reciprocity R quantified the proportion of bidirectional edges. Closeness centrality C_C measured the mean distance from each category to all others. Eigenvector centrality C_E and PageRank (α = 0.85; [17]) assessed the influence of each category weighted by the influence of its neighbors. The HITS algorithm [18] decomposed vertex importance into hub and authority scores. Directed transitivity and the mean undirected clustering coefficient quantified triangular closure. Degree assortativity r measured the correlation between the degrees of connected vertices, with negative values indicating hub-and-spoke topology [19]. Weighted and unweighted average shortest path lengths were computed.

As emphasized in Fig 1, Steps 4 and 6, the temporal sequence of clinical dialogue is implicitly encoded in all metrics because every measure is computed on the directed edges of the graph, and these edges represent aggregated temporally ordered transitions across all 110 sessions. Reciprocity is computed as the proportion of edges (u, v) for which the reverse edge (v, u) also exists. An escalation chain is identified when the network contains a directed path from a lower-severity category to a higher-severity category. Frequent transitions are those whose weight exceeds the 75th percentile of the edge weight distribution.

### Self-Loop Exclusion

As illustrated in Fig 1, Step 5, self-transitions (c_p = c_q) were systematically excluded from graph construction. This methodological choice prioritizes the analysis of dynamic category shifts and prevents prolonged exploration of a single category from dominating the network structure. The trade-off involves a loss of information about within-category dwell time in exchange for a clearer representation of transition dynamics between distinct clinical themes.

## Code Availability

Code for the construction of personalized networks from therapy sessions was adapted from Owtscharow [20], a pipeline originally developed for EEMM process-based therapy dimensions and substantially modified for the present application. The principal modifications included a backend migration from vLLM to LM Studio for privacy-preserving local inference, replacement of the EEMM classification schema with the C-SSRS categorial framework, introduction of a sliding context window (K = 8) for disambiguation, and implementation of dual-speaker classification logic.

## Statistical Analysis

Tests of Hypothesis 1(a) and 1(b) were based on descriptive comparison of therapist versus patient centrality values using Mann-Whitney U tests. Hypothesis 1(c) was tested via Spearman rank correlation between C-SSRS ideation level and closeness centrality across the eight network nodes. Hypothesis 2(b) was evaluated descriptively by comparing the mean C-SSRS ideation level of upstream versus downstream categories of ACTIVE_IDEATION. Hypothesis 3 was tested via Spearman correlation between C-SSRS ideation level of the target category and the frequency of therapist-to-patient edges (n = 6 edge types). Hypothesis 4 was tested via Spearman correlation between session-level reciprocity and the number of distinct C-SSRS categories per session, restricted to the 28 sessions with at least two suicidality-relevant utterances and at least one differential transition edge. All analyses were conducted in Python 3.12 using NetworkX and SciPy, and figures were rendered using Matplotlib [21].

### Use auf artificial intelligence, generated pre-trained transformers etc

Generative AI tools (DeepL and Claude) were used for language correction during manuscript preparation. Code review and debugging was done in Visual Code with a Claude Code plug-in. Sensitive data (on suicidality) was analysed locally with locally running models (ollama v0.33.2).

## Results

### Sample Characteristics

Complete demographic and clinical data were available from electronic health records for 106 of the 110 participants. Table 1 presents the demographic and clinical characteristics of the sample. Participants ranged in age from 19 to 65 years (M = 38.6, SD = 14.1). The sample was predominantly male (n = 57, 53.8%). The most frequent primary ICD-10 diagnosis was recurrent depressive disorder, current episode severe without psychotic symptoms (F33.2; n = 24, 22.6%), followed by single-episode severe depressive disorder (F32.2; n = 9, 8.5%) and bipolar affective disorder, current episode severe depression (F31.4; n = 5, 4.7%). Affective disorders collectively accounted for the majority of primary diagnoses. Clinical severity at intake was in the markedly ill range on the Clinical Global Impression–Severity scale (CGI-S; M = 4.7, SD = 0.9), with Global Assessment of Functioning (GAF) scores indicating serious impairment (M = 46.2, SD = 16.7). On the Beck Depression Inventory–II (BDI-II), the mean score fell within the moderate-to-severe depression range (M = 27.3, SD = 12.9; [22]), and the mean Brief Psychiatric Rating Scale (BPRS) total indicated moderate overall psychopathology (M = 42.9, SD = 10.9; [23]).

**Table 1.** Demographic and Clinical Characteristics of the Study Sample (N = 106 matched participants). CGI-S = Clinical Global Impression–Severity scale (range: 1–7). GAF = Global Assessment of Functioning (range: 1–100). BDI-II = Beck Depression Inventory–II (range: 0– 63). BPRS = Brief Psychiatric Rating Scale (18-item version; range: 18–126). Four participants could not be matched to clinical data and are therefore omitted from this table.

| Characteristic | n | % | M (SD) |
| --- | --- | --- | --- |
| Age (years) | 99 | — | 38.6 (14.1) |
| Gender | 99 |  |  |
| Male | 57 | 57.6 |  |
| Female | 41 | 41.4 |  |
| Diverse | 1 | 1.0 |  |
| Education | 96 |  |  |
| University degree | 36 | 37.5 |  |
| Vocational training | 22 | 22.9 |  |
| Upper secondary (Abitur) | 21 | 21.9 |  |
| Intermediate secondary | 11 | 11.5 |  |
| Lower secondary | 4 | 4.2 |  |
| No degree | 2 | 2.1 |  |
| CGI-S (t1) | 99 | — | 4.7 (0.9) |
| GAF (t1) | 95 | — | 46.2 (16.7) |
| BDI-II (t1) | 84 | — | 27.3 (12.9) |
| BPRS (t1) | 98 | — | 42.9 (10.9) |

### Descriptive Classification Results

Across the 110 clinical intake interviews, 506 of 55,796 utterance-level segments (0.91%) from 85 sessions (77.3%) were classified as suicidality-relevant. The remaining 25 sessions (22.7%) contained zero suicidality-relevant utterances. The mean number of suicidality-relevant utterances per session (including the 25 sessions with zero relevant utterances) was 4.60 (SD = 5.99, range: 0 to 31, median = 3.00, IQR = 0 to 6). Among the 85 sessions with at least one relevant utterance, the mean was 5.95 (SD = 6.16). Of these, 364 (71.9%) were spoken by the therapist and 142 (28.1%) by the patient. Therapist utterances were exclusively classified as THERAPIST_INQUIRY. Patient utterances spanned eight of the 11 patient-facing C-SSRS categories. ACTIVE_IDEATION (n = 67, 13.2% of all suicidality-relevant utterances) and PASSIVE_WISH (n = 33, 6.5%) were the most frequent. The categories NSSI, INTERRUPTED_ATTEMPT, PREP_BEHAVIOR, and THIRD_PERSON_MENTION were not assigned to any utterance in this corpus. Table 2 provides the full frequency distribution. Fig 2 displays the category transition network, and Fig 3 provides the ideation-level distribution.

**Table 2.**
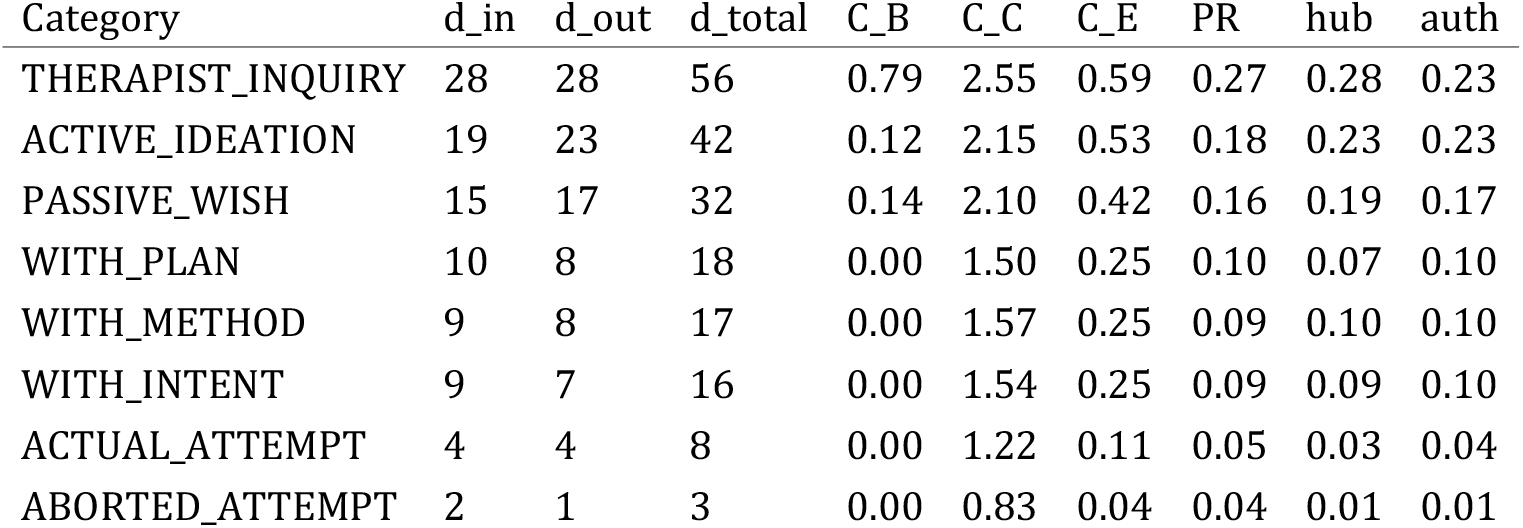
Frequency of C-SSRS Categories Across All 506 Suicidality-Relevant Utterances. Each utterance may carry multiple C-SSRS categories; the table reports the first-listed (dominant) category. Percentages are calculated **on the basis of** 506 total utterances. NSSI = non-suicidal self-injury.

**Fig 3.**
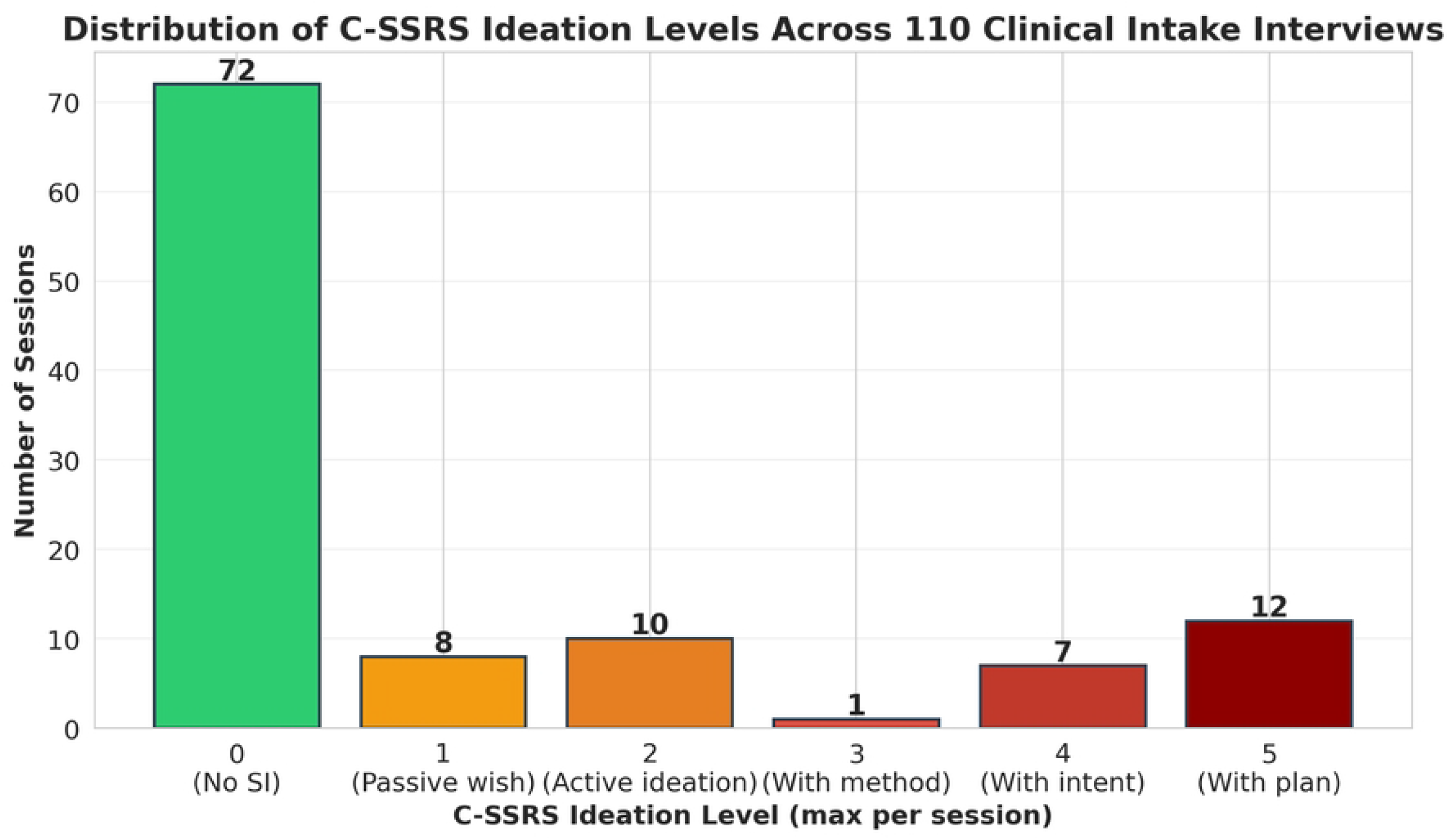
Distribution of C-SSRS ideation levels across the 110 clinical intake interviews. The ideation level for each session was derived by applying the maximum-aggregation rule of the C-SSRS to all patient self-report categories observed in that session: Level 0 = no suicidality-related utterance; Level 1 = passive death wish; Level 2 = active suicidal ideation; Level 3 = ideation with method; Level 4 = ideation with intent; Level 5 = ideation with concrete plan. Colors follow the clinical severity gradient used throughout.

### Network Structure (Hypothesis 1)

The full set of 96 differential category transitions, aggregated across all 110 sessions, yielded a directed weighted graph with |V| = 8 nodes and |E| = 32 directed edges. Four categories—NSSI, INTERRUPTED_ATTEMPT, PREP_BEHAVIOR, THIRD_PERSON_MENTION—were not detectable by the LLM. The remaining eight categories formed a densely connected directed network with density ρ = 0.57, indicating that 57.1% of all possible directed transitions between distinct categories were observable. (Hypothesis 1(a), which predicted a dominant therapist-inquiry hub, was supported. THERAPIST_INQUIRY accounted for 56 of the 96 total weighted transitions (58.3%) with perfectly symmetric in-degree and out-degree (28 each). The therapist node dominated betweenness centrality (C_B = 0.79 vs. M_patient = 0.04, Mann-Whitney U = 0.00, p = .004), confirming that nearly all shortest paths between non-therapist categories passed through the therapist node (see Table 3 for the complete centrality matrix).

**Table 3.**
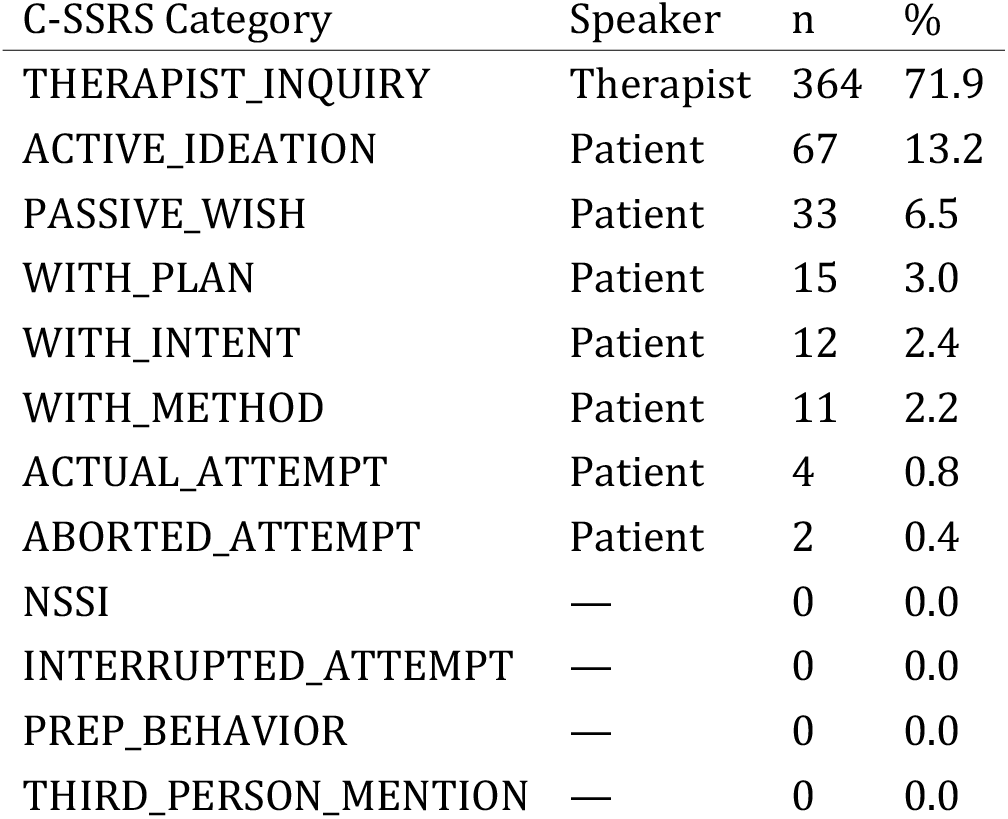
Centrality Metrics for the Eight Vertices of the Category Transition Network. **d_in** = weighted in-degree; **d_out** = weighted out-degree; **d_total** = weighted total degree; C_B = weighted betweenness centrality; C_C = weighted closeness centrality; C_E = eigenvector centrality; PR = PageRank (α = .85); hub = HITS hub score; auth = HITS authority score. Categories are sorted by **d_total** in descending order.

Hypothesis 1(b), predicting high dialogical reciprocity (R > 0.70), was supported: R = 0.81, with 26 of 32 directed edges having a reciprocal counterpart. The dominant bidirectional edge was THERAPIST_INQUIRY ↔ ACTIVE_IDEATION (w = 11 forward, w = 8 backward). Hypothesis 1(c), predicting a severity-peripherality gradient in closeness centrality, was supported: C_C decreased monotonically from THERAPIST_INQUIRY (2.55) to ABORTED_ATTEMPT (0.83), with Spearman ρ = ™.90, p = .002.

Additional structural properties were consistent with a hub-and-spoke architecture. Degree assortativity was r = ™0.23. Negative values indicate that high-degree nodes connect preferentially to low-degree nodes, the defining signature of a hub-and-spoke architecture in which a central hub links to peripheral spokes. Directed transitivity was T = 0.60. This indicates that when category A transitions to B and B transitions to C, A also tends to transition to C, and the mean undirected clustering coefficient was 0.74. The unweighted average shortest path length was 1.45, with a diameter of three steps. These values are consistent with small-world characteristics [27], though this interpretation must be made while realizing the small graph size (|V| = 8 nodes, |E| = 32 edges). Edge weights were right-skewed (M = 3.0, SD = 2.7, range: 1 to 11, median = 2.0). Notably, 9 of 32 edges (28%) carried the minimum weight of w = 1, a further 9 edges carried w = 2, and together more than half of all edges rested on one or two observed transitions.

### Bridge Function of Active Ideation (Hypothesis 2)

Hypothesis 2(a), predicting that ACTIVE_IDEATION would show the highest HITS authority score among patient categories, was partially supported. ACTIVE_IDEATION tied with THERAPIST_INQUIRY as the highest-authority node (auth = 0.23 each), reflecting that these two categories were the most frequent destinations of incoming transitions. Hypothesis 2(b), predicting that ACTIVE_IDEATION serves as a bridge to higher-severity categories, was supported. Six distinct downstream categories received transitions from ACTIVE_IDEATION, the most diversified source node among patient categories. The most frequent outgoing transitions led to WITH_METHOD (w = 4), PASSIVE_WISH (w = 6), and WITH_PLAN (w = 2). The mean C-SSRS ideation level of downstream categories (M = 3.2, SD = 1.8) was descriptively higher than that of upstream categories (M = 2.5, SD = 1.3), t(8) = 0.73, p = .49, though this difference did not reach statistical significance.

### Severity-Avoidance Gradient (Hypothesis 3)

Hypothesis 3 predicted a severity-avoidance gradient, such that therapist-initiated transitions would occur less frequently to more severe C-SSRS categories, consistent with clinician caution or conversational avoidance when probing clinically sensitive content. The six therapist-to-patient transition types and their frequencies were: THERAPIST_INQUIRY→ ACTIVE_IDEATION (w = 11), THERAPIST_INQUIRY → PASSIVE_WISH (w = 6), THERAPIST_INQUIRY → WITH_INTENT (w = 3), THERAPIST_INQUIRY → WITH_PLAN (w = 3), THERAPIST_INQUIRY → WITH_METHOD (w = 3), and THERAPIST_INQUIRY →ACTUAL_ATTEMPT (w = 2). The Spearman rank correlation between the ideation level of the target category and transition frequency was ρ = ™.66, p = .156. Although the effect was directionally consistent with the hypothesized severity-avoidance gradient, it did not reach significance. Hypothesis 3 was not supported at α = .05.

### Reciprocity and Exploration Breadth (Hypothesis 4)

Hypothesis 4 predicted that sessions with higher dialogical reciprocity would explore a greater breadth of distinct C-SSRS categories, reflecting a more thorough and bidirectional clinical suicide risk assessment. Across the 28 sessions with at least two suicidality-relevant utterances and at least one differential transition edge, the mean session-level reciprocity was 0.49 (SD = 0.46, range: 0.00–1.00). Of these, 10 sessions (35.7%) exhibited perfect reciprocity, while 12 (42.9%) exhibited zero reciprocity. The number of distinct categories per session ranged from 2 to 5 (M = 2.8, SD = 1.0). The Spearman rank correlation between session-level reciprocity and category count was ρ = .18, p = .371. Hypothesis 4 was not supported. The substantial proportion of sessions at the extremes of the reciprocity distribution suggests that session-level conversational dynamics may fall into qualitatively distinct regimes.

## Discussion

This study applied network analysis to the sequential category structure of suicide-related clinical dialogue, modeling the conversation itself as a directed weighted graph. Three findings emerged: First, suicide-related clinical communication is organized as a therapist-centered hub-and-spoke network in which the therapist functions as the dominant gateway of transitions between all C-SSRS categories (C_B = 0.79) and accounts for more than half of all observed transitions. Second, the network exhibits high reciprocity (R = 0.81) and a consistent severity-peripherality gradient in closeness centrality (ρ = ™.90), such that more clinically severe categories occupy increasingly peripheral topological positions. Third, active suicidal ideation functions as a bridge node, receiving the most transitions from therapist probes.

### Methodological Innovation

These findings should be interpreted as a proof-of-concept demonstration: the primary contribution is the methodological framework itself. It extends network theory from intra-individual symptom structures to the sequential dynamics of clinical dialogue. The primary methodological contribution of this study is the extension of network-theoretic modeling from the intra-individual domain to the inter-individual domain of clinical communication. Borsboom’s [5] network theory of mental disorders conceptualizes psychopathology as a system of mutually reinforcing symptoms; the present work demonstrates that the same formal framework can be productively applied to the sequential structure of the clinical dialogue through which those symptoms are assessed. Instead of modeling partial correlations between cross-sectional self-report items, we modeled temporally ordered transitions between categories of clinical communication, treating the conversation as a living network that unfolds in real time. This approach bridges two previously separate literatures: the symptom network tradition in psychopathology research [6,7] and the emerging field of computational analysis of clinical communication using natural language processing [11,28].

The network metrics we applied are standard tools in network science [15,16], but their application to within-session conversational dynamics is novel. Prior work applying network analysis to psychotherapy process has used Markov transition networks to characterize shifts between abstract and emotional language within sessions [9], but did not employ the full suite of graph-theoretic centrality and topology measures. The present study demonstrates that these measures yield clinically interpretable insights about the structure of suicide risk assessment dialogue, including the identification of structural bottlenecks (therapist inquiry as sole broker), bridge categories (active ideation), and topological signatures of conversational power asymmetry (hub-and-spoke architecture).

### Clinical Interpretation

The near-total dependence of the network on the therapist hub carries important clinical implications. The betweenness centrality of THERAPIST_INQUIRY (C_B = 0.79) indicates that the therapist is the central gate through which suicidality-related conversational flow passes. In the absence of therapist probes, the network would fragment into isolated nodes, a structural finding that empirically confirms the clinical axiom that suicide risk is rarely disclosed spontaneously and requires active, structured exploration [14].

The bridge function of ACTIVE_IDEATION suggests that this category represents a critical clinical juncture. When a patient discloses active suicidal ideation, the network structure indicates that the conversation is poised to transition toward more severe content.

Clinicians should recognize this disclosure not merely as a symptom to be noted, but as a conversational gateway: the moments following the disclosure of active ideation represent a structured opportunity to systematically explore escalation pathways.

The severity-peripherality gradient reveals that the most severe clinical content is also the least networked. Categories such as ACTUAL_ATTEMPT and ABORTED_ATTEMPT are structurally isolated. For any kind of AI-based automated risk monitoring systems, this finding implies that high-severity categories may appear with fewer structural precursors in the conversational network, making them harder to anticipate from preceding dialogue patterns alone. It should be noted, however, that this severity-peripherality gradient may be driven in part by the unequal frequency distribution of C-SSRS categories: the most peripheral nodes (ACTUAL_ATTEMPT, ABORTED_ATTEMPT) are also the rarest (n = 4 and n = 2 utterances, respectively), and their structural isolation may reflect limited opportunity for co-occurrence in the data rather than a genuine conversational dynamic. Disentangling frequency-driven effects from genuine topological patterns will require larger samples with greater representation of high-severity categories.

### Limitations

TSeveral limitations should be noted: The use of the first-listed category per utterance discards within-utterance category co-occurrence, which was present in 16 utterance-level pairs. emporal discounting was not applied: transitions between utterances separated by many non-suicidality-relevant utterances received equal weight to those occurring in immediate succession. The network aggregates across sessions, collapsing individual differences in conversational dynamics; multilevel network models that simultaneously estimate within-session transition structures and between-patient heterogeneity would substantially strengthen the approach [6].

A further limitation concerns the granularity at which the underlying LLM classifications have been externally validated. Independent human expert ratings were done for a subsample of n = 30 t1 transcripts from this cohort, but for practical reasons only at the whole-session level (one C-SSRS severity rating per transcript). While session-level agreement was substantial, this does not directly establish the accuracy of the individual utterance-to-category assignments that determine node membership and edge weights in the network. However, a more granular utterance-level validation is not possible for >50,000 utterances, so a session-level validation seems adequate for underscoring the plausibility of our method.

TThe absolute number of suicidality-relevant utterances (506 of 55,796; 0.91%) means that the transition network is built on a sparse empirical foundation. The study’s principal findings—the therapist-centered hub-and-spoke topology, the severity-peripherality gradient, and the bridge function of active ideation—emerged despite this sparsity and are theoretically coherent. However, their robustness and generalizability remain to be established. The severity-peripherality gradient (Spearman ρ = ™.90, p = .002) is driven by the two rarest categories occupying the most peripheral network positions; this pattern could reflect genuine conversational dynamics but could also arise from the unequal frequency distribution of categories. Future work with larger corpora drawn from settings with higher base rates of suicide-related content—such as crisis hotline transcripts, emergency department assessments, or dedicated suicide risk assessment interviews—will be essential to determine whether the network architecture described here replicates. Such larger datasets could also enable subject-specific networks, which might ultimately support individualized clinical interventions. Until such replication is available, the present findings should be regarded as a methodological proof-of-concept demonstration of a novel analytical framework rather than as definitive evidence about the structure of suicide-related clinical communication. ()

### Future Directions

First, the category transition network could be integrated with more complex symptom network models. Rather than treating C-SSRS categories as the terminal level of analysis, future work could embed them within a multilevel framework in which conversational transitions (Level 1) are nested within within-session symptom dynamics (Level 2), in turn nested within between-patient network structures (Level 3). This would allow for the simultaneous estimation of how individual differences in patients’ symptom networks shape the conversational dynamics of their clinical conversations but needs larger and more fine-grained samples. More broadly, datasets from settings with higher base rates of suicide-related content (e.g., emergency departments, crisis helplines, suicide-focused clinical trials) would enable more robust edge-weight estimation and may reveal additional network structure—such as direct transitions between high-severity patient categories— not visible in the present sparse corpus.

Second, the incorporation of therapeutic alliance measures or proxies like the Working Alliance Inventory [29], or linguistic synchrony could test whether some sessions are characterized by stronger therapeutic alliances and better clinical suicide discussions.

Third, the approach could be extended to multimodal data. The SPEAK-SAFE study [12] was designed to collect synchronized audio and transcript data, enabling the integration of acoustic features (e.g. prosody) andvisual features (facial expression, gaze direction) with the text-based category transition networks described here. Multimodal approaches to automated depression and suicide risk assessment have shown promise in systematic reviews [30,31], but have not yet been combined with network-theoretic models of conversational dynamics. A multimodal network in which prosodic markers of distress, linguistic markers of suicidality, and visual markers of affect are jointly modeled as interlocking dynamic systems would represent a significant step toward ecologically valid, clinically interpretable AI for suicide risk assessment.

## Data Availability

Due to the sensitive nature of the data and the informed consent obtained from participants, raw audio recordings and transcripts from the SPEAK-SAFE study (trial registration DRKS00027878) cannot be made publicly available. De-identified data access requests can be directed to the Ethics Committee of the University Hospital Frankfurt, which approved and over-sees the SPEAK-SAFE study, subject to a data use agreement. The code for network construction and analysis, together with the aggregated (non-identifying) category transition frequencies and network metrics underlying the reported results, is provided as Supporting Information accom-panying this article.

https://github.com/EugenOw/Create-Personalized-Networks-From-Therapy-Sessions/

## Acknowledgments

This work was supported by the ATHENE project “Privacy-Aware Domain-Adaptive Medical Natural Language Processing (PADAM)” and by the LOEWE Center DYNAMIC (Dynamic Network Approach of Mental Health to Stimulate Innovations for Change) and by a grant from the Reiss foundation (Frankfurt am Main), funded through the Hessian research funding programme LOEWE. Generative AI (DeepL and Claude) were used for language correction.

## Supplementary Material

### The following supplementary figures are available online

*S1 Fig*. Frequency of C-SSRS Categories *Across 506 Suicidality-Relevant Utterances. Categories are ordered by descending frequency. THERAPIST_INQUIRY was the most frequent (n = 364, 71*.*9%), followed by ACTIVE_IDEATION (n = 67, 13*.*2%) and PASSIVE_WISH (n = 33, 6*.*5%). Four categories in the C-SSRS schema were not observed in this corpus*.

*S2 Fig. Scatter Plot of Total Utterances per Session Against Suicidality-Relevant Utterances per Session. Each point represents one of the 110 clinical intake interviews. Point color encodes the session-level C-SSRS ideation level (0–5), with warmer colors indicating higher severity. Substantial variability is evident: some long sessions contained no suicidality-relevant utterances, while some moderate-length sessions contained up to 31*.

## Data Availability

Due to the sensitive nature of the data and the informed consent obtained from participants, raw audio recordings and transcripts from the SPEAK-SAFE study (trial registration DRKS00027878) cannot be made publicly available. De-identified data access requests can be directed to the Ethics Committee of the University Hospital Frankfurt, which approved and oversees the SPEAK-SAFE study, subject to a data use agreement. The code for network construction and analysis, together with the aggregated (non-identifying) category transition frequencies and network metrics underlying the reported results, is provided as Supporting Information accompanying this article.

## Funding

This work was supported by the ATHENE project “Privacy-Aware Domain-Adaptive Medical Natural Language Processing (PADAM)”, by the LOEWE Center DYNAMIC (Dynamic Network Approach of Mental Health to Stimulate Innovations for Change), and by a grant from the Reiss Foundation (Frankfurt am Main), funded through the Hessian research funding programme LOEWE.

## Competing Interests

Andreas Reif received speaker’s honoraria and/or served on advisory boards from Shire/Takeda, Medice, Janssen, Servier, and SAGE. Oliver Grimm received speaker’s honoraria and/or served on advisory boards from Takeda, Medice, and Boehringer Ingelheim. The other authors declare no competing interests. No author has financial relationships with any commercial entities that could directly affect or bias the results of this research. This does not alter our adherence to PLOS Digital Health policies on sharing data and materials.

## Author Contributions

Conceptualization: Oliver Grimm. Methodology: Oliver Grimm, Christopher Landau, Sofia Arampatzi. Software: Oliver Grimm, Anmol Goel, Eugen Owtscharow, Hiba Arnaout. Validation: Sofia Arampatzi, Christopher Landau. Formal Analysis: Oliver Grimm. Investigation: Christopher Landau, Sofia Arampatzi. Data Curation: Christopher Landau, Sofia Arampatzi. Writing – Original Draft: Oliver Grimm. Writing – Review & Editing: Christopher Landau, Sofia Arampatzi, Anmol Goel, Eugen Owtscharow, Hiba Arnaout, Iryna Gurevych, Andreas Reif. Visualization: Oliver Grimm. Supervision: Andreas Reif, Iryna Gurevych. Project Administration: Oliver Grimm.

## Notes

### Competing Interest Statement

I have read the journal's policy and the authors of this manuscript have the following competing interests:: Andreas Reif received speaker's honoraria and/or served on advisory boards from Shire/Takeda, Medice, Janssen, Servier, and SAGE. Oliver Grimm received speaker's honoraria and/or served on advisory boards from Takeda, Medice, and Boehringer Ingelheim. The other authors declare no competing interests. No author has financial relationships with any commercial entities that could directly affect or bias the results of this research.

### Clinical Trial

DRKS00027878

### Clinical Protocols

https://drks.de/search/en/trial/DRKS00027878

### Author Declarations

The study was approved by the institutional ethics committee (university hospital Frankfurt am Main, Germany)

